# “How about me giving blood for the COVID vaccine and not being able to get vaccinated?” A cognitive interview study on understanding of and agreement with broad consent for future use with cohort participants and guardians in Colombia and Nicaragua

**DOI:** 10.1101/2022.10.14.22281094

**Authors:** Lauren Maxwell, Jackeline Bravo Chamorro, Luz Marina Leegstra, Harold Suazo Laguna, María Consuelo Miranda Montoya

## Abstract

Informed consent (IC) is key to generating and maintaining research participants’ trust and upholding the ethical principle of autonomy. Broad consent for future use, wherein researchers ask participants for permission to share participant-level data and samples collected within the study for purposes loosely related to the study objectives, is central to enabling ethical data and sample reuse. Ensuring that participants understand broad consent-related language is key in maintaining trust in the study itself and in public health research generally. Therefore, we conducted 52 cognitive interviews with the help of semi-structured interview guides to explore cohort research participants’ understanding of the broad consent-related language in the University of California at Berkeley template IC form for biomedical research with participants from long-standing infectious disease cohort studies in Nicaragua and Colombia. After the first round of interviews, we used participants’ explanations to modify the broad consent-related language in the IC template and consequently re-evaluated participants’ understanding of and agreement with the new consent form. Participants generally supported data and sample sharing but expressed concerns about the intentions of for-profit groups as well as misuse of data or samples. Moreover, they felt uncomfortable not receiving information about incidental findings and results from future studies. Cohort participants did not understand abstract concepts including the collection and reuse of genetic data in either version of the IC. Trust in the research team and the belief that data and sample sharing could lead to new scientific insights and improved treatments were key to participant support for data and sample sharing. The study findings and research participants’ language to describe broad consent provide essential insights for researchers who want to include broad consent and ethics review committees (ERCs) working to ensure research is conducted in keeping with the ethical principle of respect for persons.

**Author Summary:** This is the first study to apply cognitive interviewing to evaluate participants’ understanding of broad consent. We found that cohort participants did not well understand the broad consent-related language in the UCB template IC. When broad consent was reworded with language from the community, difficult to process concepts, like genetic studies, were better understood. Many ethics review committees (ERCs) and universities require researchers to use a template when consenting participants for participation in biomedical research. Our finding that key concepts in the broad consent section of the template, including genetic studies and sample collection, were not well understood suggests that ERCs must allow researchers flexibility to alter IC template language to ensure participant understanding. While, like many ERCs, the University of California at Berkeley (UCB) recommends that researchers not communicate incidental findings, participants did not understand that incidental findings would not be shared and universally wanted to learn about incidental findings. Participants generally believed in the utility of data and sample reuse but expressed concerns about reuse by commercial groups and wanted to better understand who and what the future users were. Participants did not mention privacy as a concern in data and sample sharing. Informing participants about incidental findings and future uses and users is an important part of maintaining trust in data and sample sharing.

## Introduction

Research funders, regulatory agencies, and journals are increasingly advocating for or requiring that individual-level health research data are shared to maximise their utility [1, 2]. The advantages of sharing data include maximising the value of the data, improving the transparency of research, and reducing the burden and costs of unnecessary duplication of research, which is especially important for low-and-middle-income countries (LMICs), which face the competing challenges of the highest-burden of infectious disease (ID) and restricted resources for research and biobanking [3-5].

Informed consent (IC) is the process of ensuring participants’ voluntary participation in research and their understanding of the direct risks and benefits associated with participation. Broad consent for future use is consent for a loosely specified range of future research purposes that may go beyond the objectives of the original study. Participants who provide broad consent are not re-contacted to participate in these additional related studies. The consent form may or may not describe the criteria for releasing data for these future uses and which group or groups will make those decisions, i.e. the data governance framework for future use. As researchers face increased pressure to share data and specimens from health-related research, researchers and ethics review committees (ERCs) are under pressure from funders, journals, and the wider research community to include broad consent language in the study IC. Understanding how to explain data sharing to research participants was cited as a priority for future research in a recent systematic review of barriers to data sharing in LMICs and primary research with research participants and ERC members and researchers [3, 5, 6].

Understanding and addressing ethical and cultural concerns regarding data and sample sharing is central to developing values-based, ethical and appropriate IC practices. IC is critical to the ethical conduct of research (principle of respect for persons) and is vital for enrolment and continuity of participation in cohort studies [3, 7]. A commonly used and widely accepted measure of consent for data sharing is broad consent, wherein consent is obtained simultaneously for the primary study and for future use of the data and samples, including sharing with research groups external to the group that conducted the study [8]. The wording used for broad consent may explicitly state or imply that participants will not be re-contacted when sharing their data [8].

While several qualitative studies in LMICs settings have been conducted on the importance of consent for data sharing and participants’ preferred methods for obtaining consent [3, 5, 7, 9, 10], very few studies have assessed participants’ understanding of broad consent [4, 8]. One study conducted in Thailand among clinical trial participants from whom broad consent was obtained showed that they did not clearly understand data sharing and had difficulty recalling the previously shared study-related information. Broad consent for data sharing adds a layer of complexity to the consent process as it involves explaining abstract concepts to research participants [4]. Frequently, investigators do not have a clear idea of how decisions related to future use will be undertaken. Participants’ understanding of future use and incidental findings is especially important for cohort studies and perhaps even more critical for ID-focused cohort studies in LMICs, whose longevity is dependent on the continued trust of the community where the cohort is based and the local and national public health authorities who are responsible for moderating communication around IDs and seen as ultimately responsible for sharing benefits derived from data collected from their constituents.

### Research objective

This project explored research participants’ and their parents or legal guardian’s understanding of the Spanish-language translation of a template IC that includes broad consent for clinical-epidemiological and genetic data and sample sharing and the Moore Clause and is mandated by the University of California at Berkeley (UCB) for UCB-sponsored human subjects research [11]. The Moore Clause, which tells participants they will not have the right to claim compensation for commercial products or other intellectual property developed using their biospecimens from human subjects research [12], is related to a lawsuit brought by a research participant whose biopsy tissue was used without his knowledge for commercial purposes. This study provides a cross-country, cross-study comparison of how research participants understand the language around broad consent for sharing clinical and human genomic data and biospecimens, incidental findings, benefit sharing, and the Moore Clause in the UCB template.

## Materials and methods

Cognitive interviewing is a qualitative research method that explores how research participants understand specific language (e.g. survey questions, survey directions) and how that understanding varies across contexts [13]. We conducted cognitive interviews to explore research participants’ and their parents’ understanding of the broad consent-related language in the UCB Institutional Review Board (IRB) biomedical studies template consent form. We used a semi-structured interview to ascertain their agreement with the concepts covered in the broad consent section, including future use of de-identified participant-level data, genetic data, and human biospecimens. We conducted a brief demographic questionnaire to ascertain key factors associated with understanding of or agreement with broad consent-related language (age group, gender, education level, occupation). We provide the text from the UCB template for each related section in Supplementary Table S1.

After the interviews, we substituted the former template language with language and terms provided by cohort participants and their parents to better describe key concepts from the UCB informed consent form (IC). We then evaluated the research participants’ understanding of the revised IC in a new sample.

The study protocol and interview guides are available in Spanish and English on the OSF website (DOI 10.17605/OSF.IO/UXVHK). This study is reported in keeping with the Cognitive Interview Reporting Framework (CIRF) Guidelines [14], with the exclusion of guidelines specific to the evaluation of questionnaires which were not relevant for this study. Results from the thematic analysis are presented in keeping with the Standards for Reporting Qualitative Research [15]. The Spanish language version of the UCB template consent form, thinking out loud exercise, IDI guide, including the thinking aloud exercise and the Spanish language version of the UCB template consent form are included as Supplementary Text 1. This research responds to the challenges faced by researchers and ERCs who must ensure voluntary and informed participation in the research they conduct or review.

### Study population & recruitment

Participants were parents or legal guardians who gave IC for their children’s participation in dengue and Zika virus- (DENV, ZIKV) related cohort and hospital-based studies or with the children themselves if they were aged 16 or older. We included cohort participants in both Nicaragua and Colombia to compare participants’ understanding of and attitudes towards the template language across the two contexts.

In Colombia, we recruited participants from the list of parents and participants in the cohort study, “Identification of priority age groups to be vaccinated against dengue in children and adolescents”, which began in 2012. In Nicaragua, we recruited participants from a current list of parents of participants in the “Prospective hospital-based study of dengue classification, case management, and diagnosis in Nicaragua” study, assembled between 2015-2018. Inclusion criteria for the Colombia [16] and Nicaragua [17] cohort studies are described elsewhere. To capture the diversity in how cohort participants perceive broad consent language in the IC, we tried to include parents and participants of different genders and parents that had and had not completed primary school.

Given the ongoing nature of these source studies, staff were in regular communication with participants. A field interviewer from each study approached potential participants during a regularly scheduled study visit or follow-up call and described the project using the recruitment script. The interviews took place between August 2021 and April 2022, during the COVID-19 pandemic, so these contacts took place over the phone or in keeping with local directives around maintaining distance and ventilation.

Included participants were adults who could provide informed consent and had signed an IC for their child’s participation in the cohort studies mentioned above. In Colombia, we included cohort study participants aged 18 or over. In Nicaragua, we could not enrol cohort participants in this study because the cohort participants were younger at enrolment into the source cohort study, and the cohort was launched later than in Colombia. In Nicaragua, we excluded parents or guardians of participants younger than six months old or whose child weighed less than eight kilos at the time of enrolment and participants that lived outside the State of Managua from the study. Because of the risks associated with written consent during the ongoing pandemic and because research on attitudes towards language around broad consent was not considered a sensitive research topic, participants provided verbal informed consent before the interview. When parents or participants refused participation, we moved to the next randomly selected parent, legal guardian, or research participant.

### Cognitive interview

We used verbal probing and think-aloud methods in the cognitive interviews [14, 18]. Verbal probing was used to clarify participants’ understanding of complex concepts, including genetic data, de-identified data, and data and sample sharing. The think-aloud method was used to understand the respondents’ thought or decision-making process and how or why participants decided to accept or reject broad consent for future use.

Cognitive interviewing is often used to evaluate research participants’ understanding of survey questions and their responses. The classical approach to cognitive interviewing, developed by Tourangeau [19], includes how participants (1) understand the question and answers, (2) retrieve information (recall), (3) weigh different answers (estimation or judgement), and (4) arrive at their decision (response). Because we were using text from the informed consent rather than a questionnaire, participants’ recall and response processes were irrelevant to our analysis, so we used a modified version of the Tourangeau model limited to comprehension and judgement.

Before beginning the pilot, interviewers met for several weeks to practice the pre-interview “think aloud” exercise and refine the interview guide. In the interviews, we evaluated the way that participants understood certain words and phrases as well as entire sections. We used probes like “tell me more about that” to explore different interpretations of the IC. After participants explained what came to mind, we used language other than that in the IC template to describe what we saw as the intent behind the related text (e.g. samples, genetic data, incidental findings) and asked participants to describe what they then understood. We iterated this process until the participant could explain in their own words what we understood as the intended meaning of the broad consent-related language.

### Pilot phase

The study IC and cognitive interview guide, including the warm-up exercises, were pilot tested with 2-3 participants or their guardians from each of the two cohorts to ensure the cultural appropriateness of our study’s IC and interview guide language.

### Changes to cognitive interview guide after piloting

After the pilot, we shortened the cognitive interview and divided the consent and the introduction to the “think aloud” exercise and the interview into two visits to shorten the interview duration and give participants additional time to process. We also switched the focus of the mock study presented in the cognitive interview from dengue to Zika virus to prevent confusion as participants initially consented to participate in a dengue-focused cohort study.

Following the initial round of cognitive interviews conducted between March 2021 and February 2022, we used participants’ responses to revise the language in the template IC. We evaluated the community-informed IC with four additional participants in each site between March and April 2022.

### Data collection

The cognitive and semi-structured interviews were conducted in Spanish. In Nicaragua, interviews were conducted face-to-face and recorded on a digital recorder in keeping with COVID-19 public health distancing regulations. In Colombia, interviews were conducted and recorded by smartphone.

### Analysis

Recordings were transcribed verbatim, and the de-identified transcripts were uploaded to MAXQDA [20] for analysis by the study team, who is fluent in Spanish. Interviews were conducted, transcribed, and analysed on a rolling basis to allow the research team to explore interpretations of the text and themes as they emerged. At least two team members independently reviewed all transcripts, and a subset of transcripts was reviewed and evaluated by all five team members.

We used thematic analysis to explore participants’ attitudes towards data sharing based on Colaizzi’s model of descriptive phenomenology [21, 22]. During the first phase of the analysis, we developed preliminary deductive codes and definitions based on (1) the interview guide; (2) literature identified through a systematic search [23]; and (3) our own experience as researchers in public health-related data and sample sharing. We developed inductive codes by reviewing interviewer field notes, written transcripts, and memos made during the analysis process. Codes are presented in Supplementary Table S2. In the initial stage, the team read transcripts several times so that we could familiarise ourselves with the data and jointly develop the codebook. Following the joint analysis stage, to ensure reliability, JBC and LML met weekly to resolve differences in the interpretation of the cognitive interview and the application of codes and identification of emergent inductive codes grounded in the transcript data. During the same period, the entire team held weekly discussions to update and refine the codebook and clarify local expressions with the help of the field interviewers (JBC, HSL), and discuss emerging themes.

During the second phase of the analysis, the team met weekly to group the interpretations of the text and codes into meaningful themes and discuss whether these themes varied across research sites, gender, level of education, and cohort participants or their parents and guardians. We conducted weekly meetings to discuss the evolution of meaning of the cognitive interviews and the thematic analysis to ensure reliability. Throughout the project, we resolved differences of opinion in interpreting the cognitive interview and the thematic analysis of attitudes towards data and sample sharing and in how the IC should be modified through consensus.

After the first phase of interviews, we modified the broad consent section of the IC template to incorporate explanations from the community to explain complicated concepts better. Revised language was chosen through a discussion with representatives of the cohort source community and the analysis team.

### Saturation

We defined saturation [24] in relation to both the cognitive interview and the thematic analysis of participant attitudes. We stopped conducting interviews after we did not identify (1) new ways that participants understood broad consent for future use language or (2) new codes or themes in respondents’ attitudes towards broad consent. Saturation was considered both within and between sites and other subgroupings, including gender and cohort participant versus parent or legal guardian.

### Reflexivity

Participants in Colombia were interviewed by a Colombian nurse and bioethicist (JBC). In Nicaragua, participants were interviewed by an economist and expert in community-based participatory research. Additional team members come from the US (LM), Argentina (LML), and Colombia (MCMM). All team members have done or currently work in Latin America. Team members are engaged in data and sample sharing projects and have experience conducting and analysing qualitative research, including cognitive interviewing and community-based participatory research. All team members reviewed translations for fidelity to local variations in Spanish. Spanish language translations of all quotes included in this manuscript are provided in Supplementary Table S3.

### Ethics statement

The study protocol and forms were approved by the Centro de Atención y Diagnóstico de Enfermedades Infecciosas-INFOVIDA (Bucaramanga, Colombia), Instituto de Ciencias Sostenibles (Managua, Nicaragua), and Universitätsklinikum Heidelberg (Heidelberg, Germany) ethics review committees.

## Results

We conducted 24 cognitive interviews with the UCB IC template in Colombia (including four pilot interviews) and 20 interviews in Nicaragua (including three pilot interviews). After the interviews with the UCB template, four interviews were conducted with new participants using the community-informed consent language in each site. At both sites, we asked women if we could ask their male partners about their child’s participation in the study because very few men were registered as parents or legal guardians in the list of cohort participants. In these cases, the fathers reported that they were unaware of their child’s participation in the original cohort study that was the source for participants for this study. Table 1 summarises basic demographic information for study participants across sites.

**Table 1.** Participant characteristics (N=52)

|  | Colombia |  | Nicaragua |  | Total |  |
| --- | --- | --- | --- | --- | --- | --- |
|  | N | (%) | N | (%) | N | (%) |
| Broad consent language |  |  |  |  |  |  |
| UC Berkeley IC | 24 | (86) | 20 | (83) | 44 | (15) |
|  | N | (%) | N | (%) | N | (%) |
| Community-informed IC | 4 | (14) | 4 | (17) | 8 | (85) |
| Role |  |  |  |  |  |  |
| Cohort participant | 10 | (36) | 0 | (0) | 10 | (19) |
| Parent | 18 | (64) | 20 | (83) | 38 | (73) |
| Legal guardian | 0 | (0) | 4 | (17) | 4 | (8) |
| Age |  |  |  |  |  |  |
| under 25 | 7 | (25) | 0 | (0) | 7 | (13) |
| 25-34 | 4 | (14) | 3 | (13) | 7 | (13) |
| 35-45 | 9 | (32) | 14 | (58) | 23 | (44) |
| over 45 | 8 | (29) | 7 | (29) | 15 | (29) |
| Gender |  |  |  |  |  |  |
| Male | 7 | (25) | 4 | (17) | 11 | (21) |
| Female | 21 | (75) | 20 | (83) | 41 | (79) |
| Other | 0 | (0) | 0 | (0) | 0 | (0) |
| Education |  |  |  |  |  |  |
| No formal education | 0 | (0) | 1 | (5) | 1 | (2) |
| Some primary school | 1 | (4) | 2 | (8) | 3 | (6) |
| Completed primary school | 3 | (11) | 0 | (0) | 3 | (6) |
| Some high school | 4 | (14) | 7 | (29) | 11 | (21) |
| Completed high school | 8 | (29) | 8 | (33) | 16 | (31) |
| Any post-secondary education | 12 | (43) | 6 | (25) | 18 | (35) |
| Employment |  |  |  |  |  |  |
| Work inside home | 5 | (18) | 10 | (42) | 15 | (29) |
| Work outside the home | 20 | (71) | 13 | (54) | 33 | (63) |
| Student | 2 | (7) | 0 | (0) | 2 | (4) |
| Unemployed | 1 | (4) | 1 | (4) | 2 | (4) |

IC=informed consent

Out of 52 cognitive interviews, eight were conducted with men. Younger participants were more likely to have completed post-secondary education and more likely to be cohort participants than older participants. Most participants who had completed post-secondary education were younger. In Colombia, which enrolled a higher proportion of younger participants than Nicaragua, there was a higher percentage of participants who had begun or completed post-secondary education than in Nicaragua.

Challenges in understanding keywords, phrases, and concepts in the IC form were relatively consistent across participants, although there were differences in how words or concepts were understood. Variation in how participants understood the broad consent language in the UCB template IC is presented in Table 2 and described in detail below.

**Table 2.** Understanding of key concepts in the University of California Berkeley informed consent template (N=44) and revised template (N=8)

| Concept | Text from UC Berkeley PIF | Interpretation | N interpretation (N=44) | Revised text | Interpretation | N interpretation (N=8) |
| --- | --- | --- | --- | --- | --- | --- |
| Storing & sharing genetic data | We would like to have additional permission to store your child's clinical information, blood samples and DNA for future genetic studies. Genetic studies | No idea what genetic studies are | 24 | We would like to have additional permission to store your child's clinical information, blood samples and DNA for future genetic studies. Genetic studies examine the genetic information (represented by DNA) passed on from parents to children. Each person has his or her own physical characteristics and even his or her way of being thanks to the genes that are passed on from parents to children. These genes are | No idea what genetic studies are | 1 |
|  |  | Genetic studies refer to paternity tests or descendants | 7 |  | Genetic studies refer to paternity tests or descendants | 2 |
|  |  | Blood type | 5 |  | Blood type | 1 |
|  | examine the genetic information (represented by DNA) passed on from parents to children. | Genetic studies are related to other experiments that are not public health-related | 1 | organised in living beings like parts of a puzzle and the one in charge of putting the puzzle together is the DNA. A fully assembled puzzle is called a genome. Sometimes the DNA has faults, it does not do its job well and the puzzle is not well assembled. When the puzzle is not well assembled, certain diseases appear in people. That is why scientists are dedicated to analysing the DNA, to check which genes are not well organised and determine whether they can develop specific drugs for helping correct that disease. | Genetic studies are related to other experiments that are not public health-related | 0 |
|  |  | Genetic manipulation/cloning | 1 |  | Genetic manipulation/cloning | 0 |
|  |  | Genetic studies relate to the study of diseases that are transmitted from parents to children | 6 |  | Genetic studies relate to the study of diseases of diseases that are transmitted from parents to children | 0 |
|  |  | Concept understood | 0 |  | Concept understood | 4 |
| Sample | Biological samples | Samples are blood or urine that are stored for research | 18 | Blood sample |  | 0 |
|  |  | Samples are blood that are used for transfusion | 1 |  |  | 0 |
|  |  | Concept of biological samples not understood, cannot put in their own words | 11 |  |  | 0 |
|  |  | Concept understood | 14 |  | Concept understood with an additional explanation provided | 8 |

PIF=participant information form, UC=University of California

### Genetic data

The terms “genetic studies”, “genetic data”, “DNA”, and “RNA” were not well understood. When asked to put these terms in their own words, participants described “genetic” as cloning or paternity detection. Thirty one of the 44 participants across the two sites had trouble explaining what a genetic study referred to.

> *“Genetic studies? Are they going to look for genes from other people in my son’s blood? That’s more or less what I understand*.*”* (P6-ASU, Colombia, 35-45 years old, female, respondent’s parent)
>
> *“Genetic studies are the samples they ask for the parents to know if the child is theirs or not*.*”* (N20-MSMR, Nicaragua, over 45 years old, female, respondent’s parent)

Participants generally understood the language that described commercial products and that the research results could become the basis for new commercial products but did not generally agree with this idea. The phrase “diagnostic tests or therapeutic agents” was not well understood.

> *“Well, the diagnostic tests are those of the laboratory, right? I don’t understand the other*.*”* (P6 ASU, Colombia, 35-45 years old, female, respondent’s parent)

### Sample storage

The concept of storing biospecimens was generally well understood and accepted by participants. The term “biobank” was associated with “a refrigerator” and a “blood bank” to preserve and safeguard the samples—participants associated biobanking with “preserving the blood” for future research.

> *“*…*just like a blood bank, they should keep them in a warehouse, in a totally cold room…so that the samples are not damaged*.*”* (C22-DAR, Colombia, less than 25 years old, male, participant)

In a few cases, participants took storage of blood samples to mean blood donation (e.g., transfusions).

> *“*…*it is where they store blood samples or blood types when people need them, and I think that is what donations are for, and they receive them in case of an emergency”*. (N6-MPH, Nicaragua, 35-45 years old, female, respondent’s parent)
>
> *“Let’s say one bad thing is in the case that, let’s say she suffers from a chronic disease and you come and, let’s say, the institution came, and they took the sample from my daughter and the person who received that blood, got sick*.*”* (N19 JALC, Nicaragua, 35-45 years old, male, respondent’s parent)

### Confidentiality

Most participants understood the confidentiality-related procedures described in the consent form, including removing direct identifiers (e.g. name, address) and assigning a code to data and samples. Participants expressed concern that the steps taken to protect participant confidentiality could mean that clinically relevant results could not be communicated to research participants.

> *“I am worried because this tells me that all data will be erased. To a certain extent, I do not agree with that idea because it makes sense that there won’t be any negative consequences. But at least I say that if you get hit [you were infected], you should be informed*.*”* (N17-JGG, Nicaragua, over 45 years old, male, respondent’s parent)

In contrast, one participant expressed the concern that sharing detailed results from genetic research could lead to the identification of individual participants.

> *“Since it’s for future Zika research, the concern was that they would find something and say, ‘In such-and-such a person we found something in their genes!’ Or what do I know, some kind of disease and publish it and the person’s name would be revealed*.*”* (N12-MMAG, Nicaragua, 35-45 years old, female, respondent’s parent)

Another participant expressed concern that the steps taken to ensure confidentiality might not be sufficient.

> *“In some cases, I feel good and in other cases not because there is always the idea: what will happen to the sample? And if the information is really going to be confidential, what one fears the most as a parent is that something is going to happen to the child, because we are giving so much information about him*.*”* (N5-CMAZ, Nicaragua, 35-45 years old, female, respondent’s parent)

### Incidental findings

The UCB template allows research teams to choose whether or not to disclose incidental findings from research-related genetic testing but recommends that researchers not disclose these findings to research participants [11]. As such, we chose the option to not disclose incidental findings in the template version used in this study. The phrase “clinically relevant results” was well understood and associated with a diagnosis or a critical health condition.

> *“Clinically relevant is when the results are bad. When something unexpected comes out, that would be relevant to the child*.*”* (P13-LDB, Colombia, 25-34 years old, female, respondent’s parent)

Research participants generally did not understand that incidental findings from future studies that reused their data or samples would not be communicated to them. Participants expected that these findings would be shared and disagreed with the language in the template IC once it was clarified.

> *“I see it as bad that they keep that result, if in any case there is some type of disease that they discover, apart from what we are doing, because the blood sample study is not only for Zika, you are telling me that there will be other types of exams or studies that are going to be done on the samples, then if you tell me that there is not going to be any individual result or that they are going to tell the person I don’t think it’s right, in that sense I put the case when a person is a blood donor*.*”* (N17 JGG, Nicaragua, over 45 years old, male, respondent’s parent).
>
> *“I think we have the right to know if something suddenly came out of that study that could affect my son’s health*.*”* (P13-LDB, Colombia, 25-34 years old, female, respondent’s parent).
>
> *“*…*if something relevant comes out, then yes, I would like them to tell me*.*”* (P4-JS, Colombia, over 45 years old, female, respondent’s parent)
>
> *“I am aware that you are going to carry out the study. Perhaps that way they will produce a vaccine to cure Zika. But if the child has something relevant, you will not tell me and that worries me*.*”* (NPILOTO2-ESLO, Nicaragua, over 45 years old, female, respondent’s parent)
>
> *“My way of thinking was that if something abnormal happened in her blood type, it would be communicated, but you tell me that it would not be revealed. Practically, [the information] is for you, and that’s it. Is that right?”* (N3-CCBB, Nicaragua, 35-45 years old, female, respondent’s parent)
>
> *“That they are going to share my blood, and well, in actuality, I would think that would be wrong because you have to know…*.*It could be that they do something wrong or that there is some illness and you don’t know about it, and afterwards, you are sick and, and you don’t even find out. That’s why it would be bad*.*”* (P11-EYP, Colombia, 35-45 years old, female, respondent’s parent)

### Broad consent for future use

Participants understood that pseudonymised data and samples could be shared in the future with researchers involved in the study of other diseases or viruses and generally agreed with this concept. In one case, broad consent was interpreted as a way for investigators to “clean their hands” of responsibility to the research participants because, when identifying information is removed, investigators can reuse the data or sample without needing to inform the participant of the results or concerning themselves with protecting the participants’ confidentiality.

> *“Well, I already authorised them to use it for the Zika study, and I also authorised the deletion of the data once, so, well, I authorise it, right? Because it is like a form that is being filled, right? To be able to rotate it, to be able to give it to other researchers, then it is as if it is already there as if they are cleaning their hands, we eliminate it (the data). We can use that sample once they eliminate the information, to avoid a problem there, right? As such, I don’t feel bad, and I’m not going to find out and it’s normal, right? The only thing I’m going to feel is like, ok, right; it’s a study, it was what I authorised. And that’s what I’m going to take with me, and after that, I don’t know what will happen*.*”* (C22-DAR, Colombia, less than 25 years old, male, participant)

#### Information about future studies

While participants understood the phrase “can be used in this study or in other studies,” they felt that the IC did not provide sufficient information about the reuse of their data and samples. Participants expected to receive information about future studies and disagreed with the idea that they would not learn about future uses or users.

> *“I am concerned that they will not give us information, they will not give us the results, we will not know anything*.*”* (N16-MAE, Nicaragua, 25-34 years old, male, respondent’s guardian)
>
> *“Imagine, something that my son’s blood has or his DNA begins to fly around the world and without knowing to whom and for whom, well, when one gives them [the samples], let’s say to this group, well, one is confident that it is for something good, right? For an investigation about this little animal [the mosquito], the consequences that it causes. But…we are going to take [the samples] to another country, to another city or other researchers, people that we don’t know, that are not working for research or other things. And that they do it without our permission, I don’t agree*.*”* (P13-LDB, Colombia, 25-34 years old, female, respondent’s parent)

#### Fear of misuse

In some cases, the lack of information about future use was related to some participants’ concern that samples could be misused to cause harm, including the development of new viruses or for cloning.

> *“I would be a little hesitant because…look at what happened with COVID, which is said to have been something bad in laboratories, so it creates uncertainty and nerves*…*I would be worried that my son’s DNA would be used to create new viruses because, although it can be beneficial, it can also be bad because suddenly something can go wrong and it is possible that what we are currently experiencing could happen again*.*”* (C16-YMA, Colombia, less than 25 years old, female, respondent’s parent)
>
> *“*…*not [your research team] but rather other laboratories take [the sample] for other research that is not for medical purposes*.*”* (C14-DMM, Colombia, 35-45 years old, female, respondent’s parent)
>
> *“My concern is nothing more than that the samples are not occupied in what they really request*.*”* (N2-MPL, Nicaragua, 35-45 years old, female, respondent’s parent)

#### Future use as a global public good

Among the reasons given to share data and samples for future studies, interviewees mentioned the chance to contribute to improving scientific knowledge, vaccine development, and public health more generally.

> *“I decided to share data and samples. As I said before, if it were not for the fact that there were people who allowed those samples to be preserved, we would not have vaccines or medicines that are doing us good right now, so I want to be a part of that good that can be done for humanity*.*”* (C17-MSF, Colombia, over 45 years old, female, respondent’s parent)
>
> *“What I am doing and what I am authorising is because I want a conclusion to be reached as such and to help so that it really becomes an investigation that can help society. Then, yes, I would give my authorisation that the [samples] can be used again for other future investigations*.*”* (C22-DAR, Colombia, less than 25 years old, male, participant)
>
> *“I agree with sharing data and samples because it would do others good, it would grow, it would really serve the research, the effort that was made, the purpose for which we can say my son donated blood, for studies to develop vaccines. So, I would like them to do many more things, many more, for the benefit of all*.*”* (P8-IJG, Colombia, 35-45 years old, female, respondent’s parent)
>
> *“I would feel lucky if a sample, say either my daughter’s or mine, was used to combat other diseases*.*”* (N6-MPHM, Nicaragua-35-45 years old, female, respondent’s parent)

#### Sharing samples and data outside of the country

Most people supported sharing data and samples with research groups outside the country as a mechanism for expediting public health advances. Participants mentioned the COVID-19 pandemic as an example of how cross-national cooperation benefited public health through fast-tracking vaccine development.

> *“I understand that the samples are not only going to be here in the country with the research teams, but they are also going to reach other countries, where I think that different points of view and different knowledge, well, it would be good because it brings together all that knowledge of each scientist, each researcher, then it is more feasible that together they can make a vaccine, like that of COVID-19, all the countries met and then together they looked to see how to do it and managed to get a vaccine, to prevent that disease*.*”* (C16-YMA, Colombia, less than 25 years old, female, respondent’s parent)
>
> *“I feel that we are giving a grain of sand to find a solution, a vaccine…We in this country do not have as much capacity to generate a vaccine. We are not like Cuba…not like other countries directly dedicated to finding a vaccine, like right now that they are finding a vaccine for COVID*.*”* (N6-MPHM, Nicaragua, 35-45 years old, female, mother of a participant)

#### Moore Clause

The Moore Clause relates to a court case where a study participant who learned that tissues derived from his cells had been used in for-profit research and development sued the State of California to be compensated for that use [12]. The term “biological samples,” used in the Moore Clause and throughout the template IC, was clear to most participants and was generally understood as blood samples. Participants did not say they wanted to receive a financial benefit from future research but expressed concern that only a small group could monopolise the benefits from research.

> *“Let’s say I contributed my samples to obtain [the Zika vaccine]*…*it [the vaccine] is going to be worth a lot, or well, it is going to be difficult to obtain it. It is not easy to reach for some people, and all this because this discovery…belongs to a group, so they benefit economically, and well*… *they are not interested in benefiting society, which is how I think it should be. And that was the reason as I told you at the beginning, that is why I gave the sample*.*”* (C22-DAR, Colombia, less than 25 years old, male, participant)

### Benefit sharing

#### Expectation of direct benefit

Participants generally disagreed with the statement, “neither you nor the investigators will benefit directly or commercially from the research” because they associated study participation with their child’s receiving improved medical care, which may be related to their experience of receiving improved care through their participation in the cohort study.

> *“Well, that the doctors come to the house and check the child as they have done so far. Because with dengue they do it, then with Zika they should do it, right?”* (C19-MD, Colombia, 35-45 years old, female, respondent’s parent)
>
> *“The visits they do, like the ones they do now for dengue because those are good. When the child gets sick from anything, I call the lady, and the doctor comes to see the child, and they give him the medicine, which is very good*.*”* (C21-NRS, Colombia, 35-45 years old, female, respondent’s parent)
>
> *“My son is going to be more cared for by the doctors, just as they do now with dengue, the doctor comes when something happens to him. That’s good for my son too*.*”* (P7-AJS, Colombia, over 45 years old, female, respondent’s parent)
>
> *“So, they call him [the doctor] every time he [my son] is sick. [The doctors] are always looking out for my son. The doctors come when something happens to him*.*”* (P9-SV Colombia, over 45 years old, female, respondent’s parent)

When people did understand that their child would not benefit directly from the reuse of their data or samples by future studies, they disagreed with this concept.

> *“I disagree with [not receiving direct benefits]. I would feel good as long as they take the child into account to do studies, to continue looking at the diseases he has or his blood, viruses that he might have so that he will be well*.*”* (P7-AJS, Colombia, over 45 years old, female, respondent’s parent)

#### Expectation of benefit to public health

Several participants mentioned the importance of data and sample sharing to contribute to scientific or practice-related advances that will improve the lives of future generations. Some of the interviewees linked their contribution to science with vaccine development as for the COVID-19 pandemic which was ongoing at the time of the interviews.

> *“Because, well, they could suddenly bring about something good. The cure for other things that arrive in the future, another epidemic or something*.*”* (C19-MD, Colombia, 35-45 years old, female, respondent’s parent)

Participants understood the reuse of data and samples as a way to develop drugs and vaccines that could benefit humanity.

> *“New generations are coming at the global level and they will be the ones who are going to benefit from these new studies. This will be a bit of a relief because there are many diseases, not just Zika, not just dengue, there are many diseases that we do not have a cure for and it would be very good if this new generation is the beneficiary so that so many people don’t continue to die*.*”* (N4-FVMD, Nicaragua,25-34 years old, female, respondent’s parent)

#### COVID-19-related benefit sharing

Participants viewed participation in research and the reuse of that research as an important contribution to epidemic response, with several participants highlighting the public health benefits of data and sample reuse in COVID-19 response.

> *“I mean, the blood is going to be conserved, but to benefit the community, that is, to make the vaccine, for example, as we are right now with the COVID situation, we were waiting for the vaccine to free ourselves a little from that situation, so the same is with Zika*.*”* (C21-NRS, Colombia, 35-45 years old, female, respondent’s parent)
>
> *“*…*science, research should be, as such, within the public realm, right? For everyone, in other words…for the benefit of society. As is the case in what we are seeing right now [during the COVID-19 pandemic], that it is also very necessary*.*”* (C22-DAR, Colombia, less than 25 years old, male, participant)

Participants also stressed the importance of ensuring access to vaccines or other preventative or treatment measures whose production was facilitated through the reuse of data or samples generated through their participation in research.

> *“How about me giving blood for the COVID vaccine and not being able to get vaccinated?”* (C18-NAP, Colombia, 25-34 years old, female, participant)

#### Data and sample sharing with the pharmaceutical industry

Participants said that sharing data and samples with the pharmaceutical industry could help develop medicines and vaccines.

> *“I think it’s very good because that way more medicines suitable for the disease would be developed*.*”* (N14-JNP, Nicaragua, 35-45 years old, female, respondent’s mother)

Some participants saw the use of data and samples by for-profit groups as a cause for concern. They associated sample reuse by industry with a lack of benefit sharing and the reuse of samples for nefarious purposes, including the development of novel viruses which participants saw as the origin of the COVID-19 pandemic and cloning.

> *“[They are] advances for them, but that they are buying something that my daughter and I are contributing at no cost just to help the investigation and that they can come and buy without our knowledge and obtain an economic benefit, with something that we are donating without getting anything in return just to help*.*”* (N13-MNRR, Nicaragua, 35-45 years old, female, respondent’s parent)
>
> *“There, I would not be so happy because when they are shared with laboratories and they bring [outside of the country] medicine, injections, pills, etc. And they are commercial, they sell them and sometimes they are very expensive so that regular people cannot pay. Those with limited resources cannot pay, only the great millionaires that have enough money, not us poor people*.*”* (N14-JNP, Nicaragua, 35-45 years old, female, respondent’s parent)

When we explained to participants that samples could be used to produce new drugs or vaccines, they were more likely to support sample sharing. Still, they highlighted the need for equitable access and specified that new drugs or vaccines developed with their samples should be made available to research participants.

> *“Let’s say I’m going to need [the vaccine] in case I’m infected and then what am I going to do? Let’s say, I contributed my samples to obtain [the vaccine] so, what am I going to do? It’s going to cost a ton of money or, well, it’s going to be difficult to get. It won’t be easily accessible to some people, and all this because that discovery now belongs to a group [the sponsors], so they benefit financially, and well then, they would not be interested in benefiting society, which is how I think it should be. And that was the reason, as I told you at the beginning, that’s why I gave the sample. I believe that science and research should be for the benefit of everyone and should not be seen as an economic benefit. As such, because we are talking about health, we are not here to commercialise, so that is what I would like*.*”* (C22-DAR, Colombia, less than 25 years old, male, participant)

### Trust in the research team

Most participants referenced trust in the research team (UIS in Colombia, Instituto de Ciencias Sostenibles in Nicaragua) as a key driver of their decision to agree to the future use of their data and samples. This trust was based on positive experience with previous research studies and their close relationships with the cohort study staff, which relates to the long-term nature of the cohorts and their efforts to engage closely with participants. The trust was also associated with the perceived benefit of their children’s improved medical attention as cohort participants, described as more accessible access to medical appointments and closer follow-up by the medical team.

> *“Well, I trust, first in the XX university… and I know that they are not going to deliver (samples, data) to whichever research group or any laboratory because yes, they must have a project, a study, it is not all [something that happens] overnight. Rather everything is well structured, right? They are professional and very ethical*.*”* (C14-DMM, Colombia, 35-45 years old, female, respondent’s parent).
>
> *“I already participated in that dengue study…and it went well for me because [the doctors] stayed there off and on. They were calling me. They were calling people asking how they were doing and all that…So I imagine that this new stage with Zika will be the same*.*”* (C18-NAP, Colombia, 25-34 years old, female, respondent’s parent)
>
> *“Until now they have behaved very well, they come and visit me, they look out for the child. So, it must be that the people from XX university are doing something good*.*”* (C20-EM, Colombia, over 45 years old, female, respondent’s parent)

While participants expressed concerns about the possibility of misuse by future groups, this fear was balanced with their belief that the research team would not allow that to happen.

> *“That’s the problem, you don’t know if it is for [a good purpose]*…*but in any case, if it’s for the benefit of the people…if it is to get some drug, some vaccine, well, yes, I would like other people to benefit as well. Well, in truth, I have faith that you will do things well*.*”* (P6-ASU, Colombia, over 45 years old, female, respondent’s parent)

### Perception of the cognitive interview study

Most interviewees said they had gained knowledge about the data and sample sharing and consent process during the cognitive interview. Participants noted that the clarification of the IC in the cognitive interview gave them confidence in the study or helped them learn about the study.

> *“I had no idea that to carry out a study, they asked for so many permissions, such as taking the samples, storing them, to use them in the future. I had no idea because I thought that they did it without getting your permission. The fact that you are telling me that [the investigator] is going to do it for this or that, to analyse the genome, to analyse the DNA, gives me confidence*.*”* (C17-MSF, Colombia, over 45 years old, female, respondent’s parent)
>
> *“I have learned a lot I had never heard before. And I keep thinking about all that information because they should tell you when they will take your blood for those studies. I had never heard all those things*.*”* (C18-NAP, Colombia, 25-34 years old, female, participant’s mother)
>
> *“But look, at the moment, sometimes, it happens that you sign and you don’t even realise it, they can even take you to prison, because you don’t know at the moment when they are so rushed, so this becomes an experience we can learn from so that we pay attention to what we are doing*.*”* (N15-LMMU, Nicaragua, 35-45 years old, female, respondent’s parent)

### Cross group differences in the understanding and acceptance of broad consent for future use of data and samples

We did not find meaningful differences in understanding or agreement with key concepts between Colombia and Nicaragua or across the gender of participants, with the caveat that we were only able to conduct eight interviews with men. Participants who had completed high school were better able to understand and describe abstract concepts, like genetic studies, in their own words than those who had not completed high school. In Colombia, we interviewed three men and two women, aged 19-21, who were themselves research participants. These participants had all finished high school, and three had post-secondary education. Both parents or legal guardians who had given their consent for their children’s participation and cohort participants themselves supported research participation. Like their parents or legal guardians, cohort participants expressed concern about sharing data or samples with for-profit companies that they saw as potentially monopolising research data.

> *“I really would not have to complain because the only thing I’m doing is giving a contribution with a sample, helping them (*…*) that’s my contribution*…*But the researchers wouldn’t be able to get to those studies either, to those answers they want to get to, without my help and that of many others, you see?”* (C24-MSB, Colombia, less than 25 years old, male, participant)
>
> *“As I told you, I am an administrator, and I am studying administration…many times companies or especially groups, large groups, and in research there also have to be those groups, what they want to do is how to take advantage or, in such a case, be the first to arrive at an answer that no one has yet… and at the moment of having that, if, let’s say, there are several groups that have the same answers, then why do they unite themselves? To make an economic profit, then they raise prices*.*”* (C22-DAR, Colombia, less than 25 years old, male, participant)

Cohort participants also questioned whether their parents understood the information communicated in the informed consent.

> *“*…*When I participated in the study… I did not receive all this information. So, you make me think that my mom, for example, has no idea about these things. So, she just said “okay,” but because she doesn’t know anything, you understand?”* (C15-CCT, Colombia, less than 25 years old, male, participant)

#### Parental consent for sharing data or samples from their children

A few parents mentioned discussing data and sample sharing with their children.

> *“I feel like we help other people, because maybe there are people who do not allow them to take their blood sample for research. In this case, I always take the consent with her [my daughter] if she decides to give the blood because she is already a lady and she also has her voice and her vote in saying yes or no. So, I feel that we are giving a grain of sand, that support on her part so that the investigations can proceed, regardless of her data or my personal data*.*”* (N6-MPHM, Nicaragua-35-45 years old, female, respondent’s parent)
>
> *“From the time [they started] the dengue study, I consulted [my daughter]. We have always given my children a little bit [of responsibility]. We have said that they make decisions from an early age. So, I am doing this not because I decided, but because she also [decided]*.*”* (N17-JGG, Nicaragua, over 45 years old, male, respondent’s parent)

### Recommendations

#### Advice to future research participants

Most interviewees spoke about the importance of participating in research studies so that future generations could benefit from the results of these studies. Participants suggested looking ahead towards future public health advances rather than individual health or economic benefits.

> *“You shouldn’t think about the individual, but rather about the group, that we must help each other, support each other as brothers and give the opportunity to a future patient, so to speak. So that they have a medicine that can be safeguarded more quickly than in the past*.*”* (P5, Colombia, 25-34 years old, male, participant)

Participants also said that research subjects should carefully read project documents to understand the study objectives and data and sample management.

> *“That first [the participants] read [the IC] well and that they understand the information that the researchers are giving them so that everything is understood, and they respond in the best way*.*”* (C16-YMA, Colombia, less than 25 years old, female, respondent’s parent)

#### How researchers can improve broad consent

Participants said that researchers needed to provide more straightforward explanations about the research process in the IC. They said the technical language in the IC was not clarified in the written form or in the verbal explanation of the form. Some research participants mentioned that, because they had only finished primary school, they needed an explanation of the complex concepts in the IC to provide truly informed consent.

> *“I would tell them that everything they do seems very good to me and that, just like you researchers, we are people willing to collaborate so that we work together. But that things should be explained well*…*so that you can always believe them*.*”* (P3-JS, Colombia, 35-45 years old, female, respondent’s parent)

Participants also expressed concern that the level of explanation and permissions could itself create concern.

> *“Well, I don’t explain myself…excuse me for the word as a Nicaraguan, that really makes me anxious*…*That you tell me that they are going to keep it a secret, like that my daughter after that study is going to take her out, they’re going to come and kidnap her, that’s what makes me understand*.*”* (N17-JGG, Nicaragua, over 45 years old, male, respondent’s parent)
>
> *“What I don’t understand is why they ask for so many authorisations. It seems that blood samples are delicate… but because they ask for so much authorisation and why they say so many things*.*”* (C18-NAP, Colombia, 25-34 years old, female, participant)

#### Results of pilot of community-informed IC

We rewrote broad consent language in the IC template using the explanations of key phrases or concepts given by research participants (see Supplementary Text S2 for the final community-informed IC). Selected confusing phrases from the template IC and suggested language to improve understanding from community member interviews is included in Supplementary Table S4. Key differences between the community’s language and that of the UC Berkeley template included:

1. Separate sections for consent for 1) data and sample collection and preservation for the correct study; 2) data and sample preservation and sharing for future studies, including the Moore Clause; 3) use of samples to derive genetic data for use in the current and future studies;
2. The clarification of the difference between participation in research and routine clinical care;
3. Clarification of the type of sample (blood sample);
4. Simple explanation of the meaning of whole genome DNA or RNA sequencing, using a “puzzle” as a representation.

The substitution of the community’s language in the template IC and the additional clarifications resulted in a longer IC (one versus two pages in the template IC). Despite the further explanation when reviewing the community-informed broad consent language with participants, half of the eight participants did not fully understand the purpose of and procedures for genetic studies. Participants still had trouble explaining genetic data in their own words, associated study participation with their child’s receiving improved medical care and expected that the incidental findings would be shared.

## Discussion

We conducted cognitive interviews with research participants who had consented to their or their children’s participation in long-standing DENV-focused cohorts in Nicaragua and Colombia to explore their understanding of and agreement with the broad consent-related language in the UCB IRB template IC, which was required for use with human subjects research that involves genetic data and biospecimens at the time of the study [25]. The UC Berkeley template IC that we used stated that incidental findings would not be communicated and participants almost universally disagreed with this statement and expressed the desire to be informed about incidental findings. How to inform participants about incidental findings is an emerging research area that requires further research [26, 27].

Participants did not understand that they would not be informed about the future uses or users and disagreed with the idea that they would not be informed about incidental findings. They wanted to know more about the types of future users and to learn about future studies’ results. While participants felt that the UCB template did not provide enough information, participants that received the template with the community-informed language felt that there was too much information. In a few cases, the increased number of explanations in the revised template had the unintended consequence of making interview participants more worried about what could happen to their or their child’s data or samples.

Participants saw data and sample reuse as an important part of contributing to open science but felt that they did not have enough clarity on the types of future uses or of future users. As with the UCB template IC, many ICs do not specify which groups will be responsible for making decisions about the future use of data or samples [28]. Investigators often straddle the line between broad and blanket consent because they would like to maximise the utility of the data and samples and may not be able to envision the possible uses of the data or samples. In the push for open data, many prominent global health journals and funders now require the publication of study data in a repository [29, 30], resulting in a range of uses that may be more closely aligned with blanket than broad consent. Some participants expressed concerns about using data or samples for commercial purposes or by bad actors. Participants cited sample reuse for illicit purposes as a potential cause of the COVID-19 pandemic, which was ongoing during this study.

While regulatory concerns around data and sample sharing focus on the threat of reidentification [31], participants generally did not cite reidentification of themselves or their children as a concern when discussing data and sample reuse. Participants’ focus on the importance of securing benefits for the community and for science rather than concerns of reidentification suggests that benefit sharing is more important to research participants than the current focus on the perceived threat of reidentification and potential consequences thereof.

Study participants said that their primary motivations for consenting to data and sample sharing as part of their child’s participation in the original cohort study were trust in the study team related to the long-standing presence of the study staff and cohort in their community and effective benefit sharing at the individual and community or global levels. Parents felt they had better access to medical staff through participation in the study, the study communicated results to individuals and the local or national health authority, and saw the cohort as otherwise benefiting their community. They felt that sharing their data and samples could (and had) enabled faster development of vaccines, citing the research response to COVID as an example. These findings are supported by prior work that found health research participants see data and sample sharing as a way to improve their own care, improve care for people who share their condition, and as an important contribution to open science [32, 33].

Several quantitative studies have reviewed how research participants understand IC language. These studies applied a quantitative score to assess IC readability [34, 35], post-IC quizzes with research participants [36], or surveys [37] and generally found that IC language was not well understood. These findings align with previous studies in LMICs that found that complex concepts in ICFs were not well understood by research participants in Bangladesh [38], Mali, Brazil, and Oman [39], with the important caveat that these studies did not evaluate participants’ understanding of broad consent-related language.

While the concept of future use was well understood, most participants had difficulty understanding scientific or medical language in the broad consent section of the template IC, including genetic studies, biological samples, de-identified data, and incidental findings. Participants reported that the UCB IC was similar to ICs they received for other studies, which also included difficult-to-understand technical and medical language that they said was generally not clarified in either the written IC or the verbal explanation.

The collection and reuse of genetic data are sensitive and challenging to explain. Previous studies indicate that ICs for genetic research do not adequately explain what will happen with incidental findings, how samples will be managed and by whom, and the governance related to data and sample reuse [39]. IC provides potential participants with information on ethical principles of beneficence, minimising the harms and maximising the benefits of participating in research, justice, the alignment of the research with the values of the community, and engaging the community in learning about the results of the study and benefiting from the research. These studies remind us that, in addition to ensuring that participants understand broad consent for future use, the IC must appropriately address all ethical concerns.

We only identified one study that evaluated understanding of the language used to convey broad consent. That study, a textual analysis of IC forms, found that most of the 29 ICs reviewed had a high Flesch score, suggesting research participants would have had difficulty understanding the broad consent text [34]. Investigators and ERCs are in a difficult place where the need to explain complex concepts in accessible language competes with making the consent form as concise as possible. One way forward could be to reposition the consent process as an open conversation rather than a list of bullet points followed by a signature.

While participants better understood the community-informed IC, the perception that study participation results in improved care at the individual level highlights the challenge of navigating the threat of coercion and the need to engage in appropriate benefit sharing when conducting research in low-resource settings. Despite the additional explanations, the use of samples to conduct genetic studies and how genetic data are then shared was not well understood, suggesting the need for building research participants’ understanding of this concept through alternative strategies that have been successful in clarifying genetic studies and sharing genetic data, like video clips [40]. The clear preference for learning more about future users and uses, being told about incidental findings, and being able to access the results of future studies, means that cohort participants in Nicaragua and Colombia had the expectation that their shared data and samples could be linked to them so that they could be recontacted. This finding posits community preferences for learning about future studies against the pressure on research teams to anonymise rather than pseudonymise participant-level data to assuage institutional concerns around GDPR compliance.

## Strengths & limitations

This study has several strengths. This is the first study to apply cognitive interview methods to assess participants’ understanding of broad consent for future use-related language included in an IC template from a large public university that conducts a large volume of biomedical studies in high- and LMICs. While the interpretation of broad consent-related language likely varies across populations and contexts, the finding that participants did not sufficiently understand the language indicates that universities and research teams should consider using qualitative approaches to develop or evaluate the language in IC forms. This study is the first empirical research we are aware of to evaluate IC for future use and provides new insight on how this lack of understanding can relate to mistrust in the research process.

This study has some limitations. Ideally, the project interviewers would have been source community members to prevent hierarchical power dynamics during the interviews. Because the project interviewers had long-standing relationships with cohort members, they could develop a good rapport. Additionally, we could not include enough male participants to allow a cross-gender comparison of responses. In both settings in Nicaragua and Colombia, women are generally responsible for taking care of educational and health-related issues for their children. This is supported by our finding that women were most often responsible for consenting to their children’s participation in the cohort studies.

## Recommendations for research and practice

The ethics community has moved away from the paradigm of ERCs in high-resourced settings evaluating the ethics of research conducted in low-resource settings to mandating in-country ethics review for multi-country studies. While this move ensures that local values that shape interpretations of shared ethical principles are included in the IC language and structure, local ERCs may be reluctant to change the language of template ICs from ERCs that manage multi-site studies and are often based in high-income settings. A 2012 qualitative study with 46 representatives from 34 US IRBs found that IRBs report several challenges in conducting ethical, culturally relevant research in LMICs. These included a lack of knowledge of local context and how the ethical principles of autonomy, justice, and proportionality are considered in different contexts [41]. The same study found that US-based ERCs’ fear of “worse contributions scenarios” related to research management sometimes led them to paternalistic approaches to interaction with ERCs in LMICs [41]. While ERCs need to ensure continuity in cross-study, cross-setting adherence to widely accepted ethical principles, context is important, and ERCs should allow investigators, which need to include representatives from the community participating in the study, sufficient leeway to tailor the consent language and content to reflect community values and preferences.

## Conclusion

This study adds to the growing body of evidence suggesting that IC is not well understood and contributes to the very limited research on how broad consent is understood. We found that participants’ trust in the research team, rather than the explanation provided in the IC, was the most important foundation for their support for data and sample sharing. While participants were concerned about sharing data and samples with for-profit groups, they generally expected that their data and samples would be reused and wanted to learn about incidental findings. Researchers should consider sharing pseudonymised rather than anonymised data to allow for the communication of incidental findings and ensure participants can access the results of future studies that use their data or samples. Cognitive interviews can help ensure that language around broad consent for future use of data and samples is well understood. In addition to meeting the ethical principle of respect for persons, providing understanding fosters trust in the research process and data or sample sharing more broadly.

## Data Availability

The study protocol and interview guides are available in Spanish and English on the OSF website (DOI 10.17605/OSF.IO/UXVHK). The de-identified cognitive interview study transcripts were uploaded to openICPSR (https://doi.org/10.3886/E173322V2).

https://osf.io/uxvhk/

## Acknowledgments

Above all, we would like to thank the Paediatric Dengue Hospital-Based Study participants of Nicaragua and the AEDES Cohort study participants of Piedecuesta, Colombia and their families for sharing their insights and time in support of this study. We would also like to acknowledge Fundación INFOVIDA and Centro de Atención y Diagnóstico de Enfermedades Infecciosas (CDI) Bucaramanga, for their support in facilitating the implementation of the study in Colombia. We are grateful to past and present study team members based at Hospital Infantil Manuel de Jesús Rivera, the National Virology Laboratory in the Centro Nacional de Diagnóstico y Referencia, and the Sustainable Sciences Institute in Nicaragua.

We also thank Dr. Priscilla Cesar for her thoughtful comments on the manuscript.

## Supplementary information

**S1 Table. Relevant sections of the University of California Berkeley informed consent**

**S2 Table. Codebook**

**S3 Table. Supporting quotes in original Spanish and English translation**

**S4 Table. Confusing phrases in template informed consent and suggested language**

**S1 Text. Interview guide that evaluates understanding of language from the Spanish translation of the University of California at Berkeley Spanish-language template informed consent for biomedical studies**

**S2 Text. Interview guide that evaluates understanding of language from the Spanish translation of the University of California at Berkeley Spanish-language template informed consent for biomedical studies updated with language from explanations from research participants**

## References

1. Taichman DB, Sahni P, Pinborg A, Peiperl L, Laine C, James A. Data sharing statements for clinical trials: a requirement of the International Committee of Medical Journal Editors. Lancet. 2017;389. doi: 10.1016/s0140-6736(17)31282-5.

2. Phillips M. International data-sharing norms: from the OECD to the General Data Protection Regulation (GDPR). Human Genetics. 2018;137(8):575–82. doi: 10.1007/s00439-018-1919-7.

3. Denny SG, Silaigwana B, Wassenaar D, Bull S, Parker M. Developing Ethical Practices for Public Health Research Data Sharing in South Africa:The Views and Experiences From a Diverse Sample of Research Stakeholders. Journal of Empirical Research on Human Research Ethics. 2015;10(3):290–301. doi: 10.1177/1556264615592386. PubMed PMID: 26297750.

4. Cheah PY, Jatupornpimol N, Hanboonkunupakarn B, Khirikoekkong N, Jittamala P, Pukrittayakamee S. Challenges arising when seeking broad consent for health research data sharing: a qualitative study of perspectives in Thailand. BMC Med Ethics. 2018;19. doi: 10.1186/s12910-018-0326-x.

5. Cheah PY, Tangseefa D, Somsaman A, Chunsuttiwat T, Nosten F, Day NP. Perceived benefits, harms, and views about how to share data responsibly: a qualitative study of experiences with and attitudes toward data sharing among research staff and community representatives in Thailand. J Empir Res Hum Res Ethics. 2015;10. doi: 10.1177/1556264615592388.

6. Bull S, Cheah PY, Denny S, Jao I, Marsh V, Merson L. Best practices for ethical sharing of individual-level health research data from low- and middle-income settings. J Empir Res Hum Res Ethics. 2015;10. doi: 10.1177/1556264615594606.

7. Hate K, Meherally S, Shah More N, Jayaraman A, Bull S, Parker M, et al. Sweat, Skepticism, and Uncharted Territory:A Qualitative Study of Opinions on Data Sharing Among Public Health Researchers and Research Participants in Mumbai, India. Journal of Empirical Research on Human Research Ethics. 2015;10(3):239–50. doi: 10.1177/1556264615592383. PubMed PMID: 26297746.

8. Mweemba O, Musuku J, Mayosi BM, Parker M, Rutakumwa R, Seeley J, et al. Use of broad consent and related procedures in genomics research: Perspectives from research participants in the Genetics of Rheumatic Heart Disease (RHDGen) study in a University Teaching Hospital in Zambia. Global Bioethics. 2019:1–16. doi: 10.1080/11287462.2019.1592868.

9. Merson L, Phong TV, Nhan LNT, Dung NT, Ngan TTD, Kinh NV, et al. Trust, Respect, and Reciprocity:Informing Culturally Appropriate Data-Sharing Practice in Vietnam. Journal of Empirical Research on Human Research Ethics. 2015;10(3):251–63. doi: 10.1177/1556264615592387. PubMed PMID: 26297747.

10. Jao I, Kombe F, Mwalukore S, Bull S, Parker M, Kamuya D, et al. Involving Research Stakeholders in Developing Policy on Sharing Public Health Research Data in Kenya:Views on Fair Process for Informed Consent, Access Oversight, and Community Engagement. Journal of Empirical Research on Human Research Ethics. 2015;10(3):264–77. doi: 10.1177/1556264615592385. PubMed PMID: 26297748.

11. Template Consent Form - Biomedical Study Berkeley, California: University of California at Berkeley; [cited 2022 March 31]. Available from: https://cphs.berkeley.edu/CF-Template_Biomed.docx.

12. Hall J. Use of Specimens (Moore Clause) Disclosure in the Informed Consent Form: University of California; 2014 [updated August 21, 2014; cited 2022 March 31]. Guidance Memo 14-07:[Available from: https://researchmemos.ucop.edu/php-app/index.php/site/document?memo=UlBBQy0xNy0wNA==&doc=3709.

13. Meadows K. Cognitive Interviewing Methodologies. 2021;30(4):375–9. doi: 10.1177/10547738211014099. PubMed PMID: 33998325.

14. Boeije H, Willis G. The Cognitive Interviewing Reporting Framework (CIRF). 2013;9(3):87–95. doi: 10.1027/1614-2241/a000075.

15. O’Brien BC, Harris IB, Beckman TJ, Reed DA, Cook DA. Standards for Reporting Qualitative Research: A Synthesis of Recommendations. Academic Medicine. 2014;89(9):1245–51. doi: 10.1097/acm.0000000000000388. PubMed PMID: 00001888-201409000-00021.

16. Estupiñán Cárdenas MI, Herrera VM, Miranda Montoya MC, Lozano Parra A, Zaraza Moncayo ZM, Flórez García JP, et al. Heterogeneity of dengue transmission in an endemic area of Colombia. PLOS Neglected Tropical Diseases. 2020;14(9):e0008122. doi: 10.1371/journal.pntd.0008122.

17. Andrade DV, Katzelnick LC, Widman DG, Balmaseda A, M. de Silva A, Baric RS, et al. Analysis of Individuals from a Dengue-Endemic Region Helps Define the Footprint and Repertoire of Antibodies Targeting Dengue Virus 3 Type-Specific Epitopes. mBio. 2017;8(5):e01205–17. doi: 10.1128/mBio.01205-17.

18. Knafl K, Deatrick J, Gallo A, Holcombe G, Bakitas M, Dixon J, et al. The analysis and interpretation of cognitive interviews for instrument development. Res Nurs Health. 2007;30(2):224-34. Epub 2007/03/24. doi: 10.1002/nur.20195. PubMed PMID: 17380524.

19. Tourangeau R, Rasinski K. Cognitive processes underlying context efects in attitude measurement. Psychol Bull. 1988;10(3):299–314. doi: https://doi.org/10.1037/0033-2909.103.3.299.

20. Plus M. MAXQDA. Berlin, Germany: VERBI Software GmbH; 2020.

21. Fisher LD, Lin DY. TIME-DEPENDENT COVARIATES IN THE COX PROPORTIONAL-HAZARDS REGRESSION MODEL. Annual Review of Public Health. 1999;20(1):145–57. doi: 10.1146/annurev.publhealth.20.1.145. PubMed PMID: 10352854.

22. Colaizzi PF. Psychological research as the phenomenologist views it. In: Valle RS, King M, editors. Existential-Phenomenological Alternatives for Psychology: Oxford University Press; 1978.

23. Maxwell L, Gilyan R, Chavan SA, Merson L, Saxena A, Terry R. Guidance for ensuring fair and ethical broad consent for future use. A scoping review protocol. F1000Research. 2021;10:102-. doi: 10.12688/f1000research.51312.1. PubMed PMID: 33953907.

24. Hennink MM, Kaiser BN, Marconi VC. Code Saturation Versus Meaning Saturation:How Many Interviews Are Enough? Qualitative Health Research. 2017;27(4):591–608. doi: 10.1177/1049732316665344. PubMed PMID: 27670770.

25. Informed Consent Berkeley, CA: UC Berkeley Human Research Protection Program; 2020 [cited 2020 January 1]. Available from: https://cphs.berkeley.edu/informedconsent.html.

26. Christenhusz GM, Devriendt K, Dierickx K. To tell or not to tell? A systematic review of ethical reflections on incidental findings arising in genetics contexts. European Journal of Human Genetics. 2013;21(3):248–55. doi: 10.1038/ejhg.2012.130.

27. Sapp JC, Facio FM, Cooper D, Lewis KL, Modlin E, van der Wees P, et al. A systematic literature review of disclosure practices and reported outcomes for medically actionable genomic secondary findings. Genetics in Medicine. 2021;23(12):2260–9. doi: 10.1038/s41436-021-01295-7.

28. Munung NS, Marshall P, Campbell M, Littler K, Masiye F, Ouwe-Missi-Oukem-Boyer O, et al. Obtaining informed consent for genomics research in Africa: analysis of H3Africa consent documents. 2016;42(2):132–7. doi: 10.1136/medethics-2015-102796 %J Journal of Medical Ethics.

29. Rothstein MA. Informed Consent for Secondary Research under the New NIH Data Sharing Policy. Journal of Law, Medicine & Ethics. 2021;49(3):489-94. Epub 2021/10/19. doi: 10.1017/jme.2021.69.

30. Christensen G, Dafoe A, Miguel E, Moore DA, Rose AK. A study of the impact of data sharing on article citations using journal policies as a natural experiment. PLOS ONE. 2019;14(12):e0225883. doi: 10.1371/journal.pone.0225883.

31. Scheibner J, Ienca M, Kechagia S, Troncoso-Pastoriza JR, Raisaro JL, Hubaux J-P, et al. Data protection and ethics requirements for multisite research with health data: a comparative examination of legislative governance frameworks and the role of data protection technologies†. Journal of Law and the Biosciences. 2020;7(1). doi: 10.1093/jlb/lsaa010.

32. Wai CT, Mackey SJ, Hegney DG. Patients’ experiences towards the donation of their residual biological samples and the impact of these experiences on the type of consent given for secondary use: A systematic review. 2011;9(42):1714–81. doi: 10.11124/jbisrir-2011-108. PubMed PMID: 01583928-201109420-00001.

33. Shabani M, Bezuidenhout L, Borry P. Attitudes of research participants and the general public towards genomic data sharing: a systematic literature review. Expert Review of Molecular Diagnostics. 2014;14(8):1053–65. doi: 10.1586/14737159.2014.961917.

34. Jacquier E, Laurent-Puig P, Badoual C, Burgun A, Mamzer M-F. Facing new challenges to informed consent processes in the context of translational research: the case in CARPEM consortium. BMC Medical Ethics. 2021;22(1):21. doi: 10.1186/s12910-021-00592-9.

35. Bothun LS, Feeder SE, Poland GA. Readability of Participant Informed Consent Forms and Informational Documents: From Phase 3 COVID-19 Vaccine Clinical Trials in the United States. Mayo Clinic Proceedings. 2021;96(8):2095–101. doi: 10.1016/j.mayocp.2021.05.025.

36. Manta CJ, Ortiz J, Moulton BW, Sonnad SS. From the Patient Perspective, Consent Forms Fall Short of Providing Information to Guide Decision Making. J Patient Saf. 2021;17(3):e149–e54. doi: 10.1097/PTS.0000000000000310. PubMed PMID: 27490160.

37. Hyun Song J, Yoon HS, Hoon Min B, Haeng Lee J, Kim YH. Acceptance and Understanding of the Informed Consent Procedure Prior to Gastrointestinal Endoscopy by Patients: A Single-Center Experience in Korea FAU - Song, Ji Hyun FAU - Yoon, Hwan Sik FAU - Min, Byung Hoon FAU - Lee, Jun Haeng FAU - Kim, Young Ho FAU - Chang, Dong Kyung FAU - Son, Hee Jung FAU - Rhee, Poong Lyul FAU - Rhee, Jong Chul FAU - Kim, Jae J. Korean J Intern Med. 2010;25(1):36–43. doi: 10.3904/kjim.2010.25.1.36.

38. Das D, Cheah PY, Akter F, Paul D, Islam A, Sayeed AA. Participants’ perceptions and understanding of a malaria clinical trial in Bangladesh. Malar J. 2014;13. doi: 10.1186/1475-2875-13-217.

39. Al-Riyami A, Jaju D, Jaju S, Silverman H. The adequacy of informed consent forms in genetic research in Oman: A pilot study. Developing World Bioethics. 2011;11(2):57–62. doi: https://doi.org/10.1111/j.1471-8847.2010.00293.x.

40. Rego S, Grove ME, Cho MK, Ormond KE. Informed consent in the genomics era. Cold Spring Harb Perspect Med. 2020;10(8):a036582. doi: 10.1101/cshperspect.a036582.

41. Klitzman RL. US IRBs Confronting Research in the Developing World. Developing World Bioethics. 2012;12(2):63–73. doi: https://doi.org/10.1111/j.1471-8847.2012.00324.x.

